# The association between lymphocyte mitochondrial DNA abundance and Stroke: a combination of multivariable-adjusted survival and mendelian randomization analyses

**DOI:** 10.1101/2021.10.27.21265463

**Authors:** Leon G Martens, Jiao Luo, Marieke J H Wermer, Ko Willems van Dijk, Sara Hägg, Felix Grassmann, Raymond Noordam, Diana van Heemst

## Abstract

**Background and Purpose:** Mitochondrial dysfunction is associated with increased Reactive Oxygen Species that are thought to drive risk of disease, including stroke. We investigated the association between mtDNA abundance, as a proxy for mitochondrial function, and incident stroke using multivariable-adjusted survival and Mendelian Randomization (MR) analyses.

**Methods:** Cox-proportional hazard model analyses were conducted to assess the association between lymphocyte mtDNA abundance, and incident ischemic and hemorrhagic stroke over a maximum of 14-years follow-up in unrelated European-ancestry participants from UK Biobank. MR was conducted using independent (R^2^<0.001) lead variants for lymphocyte mtDNA abundance (p < 5×10-8) as instrumental variables. Single-Nucleotide Polymorphism (SNP)- ischemic stroke associations were derived from three published open source European-ancestry results databases (cases/controls): MEGASTROKE (60,341/454,450), UK Biobank (2,404/368,771) and FinnGen (10,551/202,223). MR was performed per study, and results were subsequently meta-analyzed.

**Results:** A total of 288,572 unrelated participants (46% men) with mean (SD) age of 57 (8) years were included in the cox-proportional hazard analyses. After correction for considered confounders (BMI, hypertension, cholesterol, T2D), no association was found between mtDNA abundance and ischemic or hemorrhagic stroke (lowest 20% versus highest 20%: ischemic stroke, hazard ratio, 1.06 [95% confidence interval 0.95, 1.18]; hemorrhagic stroke, hazard ratio 0.97 [95% confidence interval, 0.82, 1.15]). In line, in the MR analyses, we found no evidence for an association between genetically-influenced mtDNA abundance and ischemic stroke (odds ratio, 1.04; confidence interval, 0.95, 1.15).

**Conclusions:** From the multivariable-adjusted survival analyses and the MR analyses, we did not find support for low lymphocyte mtDNA abundance as a causal risk factor in the development of stroke.

## Introduction

Stroke is the second leading cause of death and loss of disability-adjusted life years worldwide.[1] Several studies have shown that oxidative stress plays an important role in the pathophysiology of stroke. Oxidative stress is known to aggravate secondary damage in both hemorrhagic and ischemic stroke and increases reperfusion injury after ischemic stroke. [2-4]. As a result of direct or indirect Reactive Oxygen Species (ROS)-induced damage to the vascular wall, multiple aspects of the vascular system are altered including platelet aggregation, endothelial function, vascular permeability and vasodilation.[3] Collectively, these alterations are detrimental to the vasculature and lead to local lesions in the cerebral vessels. [3] These local vessel changes induced by oxidative stress can also gradually develop before the occurrence of stroke. Therefore oxidative stress via an excess of ROS may not only play a pivotal role in secondary damage after stroke but may also lead to an increased risk of stroke incidence. [5]

Mitochondria are a major source of ROS production.[6] Mitochondrial dysfunction leads to an increase in ROS production due to a change in redox homeostasis.[7] The mitochondrial DNA copy number (mtDNA-CN) is seen as a proxy of mitochondrial function.[8, 9] mtDNA-CN in lymphocytes can be assessed relatively easy by estimating the mtDNA abundance from the intensities of genotyping probes representing mitochondrial DNA on genotyping arrays.[10]

Due to the extraordinary energy demand of the brain, which is produced by the mitochondria, there is an increased amount of ROS production.[11] Given the postulated detrimental biological effect of oxidative stress on the cerebrovascular system and its role in secondary damage after stroke occurrence, we hypothesized that lower mtDNA-CN could also be associated with a higher risk of stroke. We, therefore, investigated the association between lymphocyte mtDNA abundance and incident ischemic and hemorrhagic stroke in a large cohort of European-ancestry participants from the UK Biobank. To investigate causality, we applied Mendelian Randomization (MR) analyses, [12, 13]. To obtain convincing evidence for a possible causal association, we set out to use two separate analysis methods, both with different assumptions and limitations, in a large study sample.[12]

## Materials and Methods

### Population description

The UK Biobank cohort is a prospective general population cohort with 502,628 participants between the age of 40 and 70 years recruited from the general population between 2006 and 2010 [14] (more information can be found online https://www.ukbiobank.ac.uk). Blood samples were collected for genotyping. Access for information to invite participants was approved by the Patient Information Advisory Group (PIAG) form England and Wales. All participants in the UK Biobank provided a written informed consent. The present study was accepted under project number 56340.

In the present study, genotyped European-ancestry participants were followed (N = 488 377). Exclusion criteria included: 1) non-European ancestry; 2) participants who failed genotyping quality control and/or with low call rate; 3) related individuals defined by principal components (PCs); 4) participants with high SD of autosomal probes; 5) history of any stroke at baseline; 6) missingness on covariates. After the exclusions, the final analyses were performed in 288,572 participants.

### Mitochondrial DNA copy numbers

We used somatic mtDNA abundance as a proxy of mtDNA-CN, as the exposure, which is determined from the intensities of genotyping probes on the mitochondrial chromosome on the Affymetrix Array. The method for computing mtDNA abundance has been described previously.[10] In brief, the relative amount of mtDNA hybridized to the array at each probe was the log2 transformed ratio (L2R) of the observed genotyping probe intensity divided by the intensity at the same probe observed in a set of reference samples. We used the median L2R values across all 265 variants passing quality control on the MT chromosome as an initial raw measure of mtDNA abundance. To correct for confounding induced by poorly performing probes, we weighted L2R values of each probe by multiplying the weight of the probe that are generated from a multivariate linear regression model in which those intensities statistically significantly predicted normalized mitochondrial coverage from exome sequencing data, resulting in a single mtDNA abundance estimate for each individual. To eliminate the plate effect, we subsequently normalized the abundance to mean of zero and SD of one within each genotyping plate consisting of 96 wells. [9]

### Covariates

Lifestyle data and medical history was provided by the participants via touch-screen questionnaires (smoking, alcohol consumption, disease status, medication use) and physical measurements (BMI, blood pressure).

### Outcome

For the outcome we looked at any stroke, as well as ischemic and hemorrhagic stroke separately, in the time period August 2006 up to January 2021. Stroke incidence was obtained via hospital admission data and national health register data and used to identify the date of the first stroke or stroke-related death after baseline assessment. The primary outcomes were any stroke incidence and further specified ischemic and hemorrhagic stroke incidence. Incident disease diagnoses are coded according to the International Classification of Diseases edition 10 (ICD-10); Stroke cases are defined as I64, ischemic stroke as I63.9 and hemorrhagic stroke as I61.9. Follow-up time is computed from the baseline visit to the diagnosis of incident disease, loss-to-follow-up or death, or the end of the study period, whichever came first.

### Data required for the Mendelian Randomization analyses

For the MR, genetic variants of mtDNA abundance were used as instrument variables. In a previous study, 129 independent SNPs as genetic variants were found to be associated with mtDNA abundance.[15] The study was performed in a total of 465,809 individuals using a combined population of the Cohorts for Heart and Aging Research in Genomic Epidemiology (CHARGE) consortium and the UK Biobank. In order to minimize potential weak instrument bias, we considered an F-statistic of at least 10 as sufficient for performing an MR analysis, which is well-accepted in the field. [16]

### Sensitivity analysis using different set of genetic instruments

As the genetic instruments from the main analysis were identified in two relatively different cohorts, the MR analysis were repeated with a second set of genetic variants from a different GWAS performed only in the UK Biobank. Therefore, 66 independent variants from 50 genomic regions were identified in a GWAS performed on mtDNA abundance using a population of 295,150.[10]

### Mendelian Randomization outcome datasets

For the extraction of summary statistics on the associations of the mtDNA abundance related SNPs with ischemic stroke, three large studies were used: the MEGASTROKE consortium, the UK Biobank, and the FinnGen study.[14, 17] Both UK Biobank and FinnGen were not part of the main analyses of the MEGASTROKE consortium preventing inclusion of overlapping samples in the analyses. In the three studies insufficient data on hemorrhagic stroke was available.

The MEGASTROKE consortium was used to retrieve the ischemic stroke outcome data and consisted of 60,341 cases and 454,450 controls collected from 29 studies of predominantly the European ancestry (86%).[17] Both UK Biobank and FinnGen were not part of the main analyses of the MEGASTROKE consortium preventing inclusion of overlapping samples in the analyses.

Different from the survival analyses, for the MR analyses on the UK Biobank cohort, any event of ischemic stroke is considered (before and after enrollment into the study). The MR analyses were performed with participants of European ancestry, who were in the full released imputed genomics databases (UK10K + HRC). Follow-up information that included ischemic stroke occurrence was retrieved through the routinely available NHS database. In the current dataset, we had data available on 2,404 cases of ischemic stroke, and 368,771 controls. We performed new genome-wide association analyses using linear mixed models to assess the associations between genetic instruments and ischemic stroke, adjusted for age, sex and 10 principal components, and corrected for familial relationships using BOLT_LMM (v2.3.2).

The FinnGen study is an ongoing cohort study launched in 2017 and used genetic data generated from biobank samples and health-related data from social and healthcare registers and combined these. Detailed information such as participating biobanks/cohorts, genotyping and data analysis are available at their website (https://www.finngen.fi/en/). For our current study, the freeze 5 data was used. Within this data, there are 10,551 reported cases of ischemic stroke, and 202,223 controls.

## Statistical analysis

### Multivariable-adjusted analyses

For the analyses, and for presentation purposes only, we divided the study population in 5 equally-sized groups based on the mtDNA copy numbers, with the first quintile containing the group with the lowest levels of mtDNA copy numbers and the fifth quintile containing the highest levels (used as reference).

Baseline characteristics of the study population were presented separately for these quintiles, as mean (SD) for continuous variables and frequency (proportion) for categoric variables.

The cumulative incidence for competing risk (CICR) was used to plot the cumulative incidence of ischemic and hemorrhagic stroke separately using a Kaplan-Meier survival curve by mtDNA abundance quintiles. For any, ischemic and hemorrhagic stroke a Cox proportional hazards model was used to estimate the hazard ratio (HR) and 95% confidence interval (CI) on incident disease risk of the mtDNA abundance quintile compared with the reference. Analyses were additionally done stratified by sex. Two multivariate regression models were fitted, where for model 2 covariates were first added individually:

- Model 1: age, sex, batch, the first two PCs, white blood cell counts, platelet
- Model 2: Model 1 + BMI, smoking, alcohol consumption, total cholesterol, hypertension, diabetes, cholesterol lowering medication, blood pressure lowering medication

Data was presented as hazard ratios and their corresponding 95% confidence interval (CI) on stroke incidence, comparing the lowest 20% mtDNA copy numbers with the highest 20%. Analyses were performed using the “Survival” (cran.r-project.org/web/packages/survival) package in R (v4.1.0)

### Mendelian Randomization

All the analyses were done using R (v4.1.0) statistical software (The R Foundation for Statistical Computing, Vienna, Austria). MR analyses were performed using the R-based package “TwoSampleMR” (https://mrcieu.github.io/TwoSampleMR/).[18]

For our primary MR analysis, Inverse-Variance weighted (IVW) regression analyses were performed. [13] Estimates were calculated for each genetic instrument using the Wald ratio (SNP – outcome association divided by the SNP – exposure association) and subsequently meta-analyzed using the inverse-weighted meta-analyses weighted on the standard error of the SNP-outcome association (assuming no measurement error [NOME] in the exposure).[19] The calculated estimates were expressed as odds ratios (OR) on ischemic stroke per SD difference in lymphocyte mtDNA abundance. However, this method assumes that none of the included genetic instrumental variables are pleiotropic, which is hardly the case when including large numbers of genetic variants as instrumental variables.

To ensure that the results obtained from the IVW analyses were not biased due to directional pleiotropy, we performed MR-Egger regression analysis and Weighted-Median Estimator.[19] Although MR-Egger is considered as a relatively inefficient approach (e.g., large confidence intervals), this method does not force the regression line to go through the intercept. The intercept depicts the estimated average pleiotropic effect across the genetic variants, and a value that differs from zero indicates that the IVW estimate is biased [20]. The Weighted-Median estimator analysis can provide a consistent valid estimate if at least half of the instrumental variables are valid. [21] In addition, MR-PRESSO (MR Pleiotropy RESidual Sum and Outlier) was applied to detect and correct for horizontal pleiotropy through removing outlying causal estimates based on individual instruments [22], as implemented in the R-based package MR-PRESSO (https://github.com/rondolab/MR-PRESSO)*”*. The Cochran’s Q statistic was performed in order to test the heterogeneity between the estimated Wald ratios from different genetic variants. [23] A power calculation was performed (https://shiny.cnsgenomics.com/mRnd/). With power = 0.80, minimal effect size (OR) was 1.076.

Recent research has proven that two-sample MR methods can safely be used for one-sample MR in large databases.[24] This allows us to use the UK Biobank database in our sample set despite also being used as our exposure dataset. As a limitation to this method, results of MR-Egger analyses are to be interpreted with caution when used to check for pleiotropy.[24]

The main MR analyses were performed in the individual datasets, and subsequently meta-analyzed to derive the pooled estimates for the exposure on the risk of ischemic stroke. Heterogeneity of the estimates across three datasets was performed by I^2^, and corresponding p-value was obtained from the Cochran’s Q test. When no heterogeneity was found amongst the three cohorts, a fixed-effect model meta-analysis was used to pool the instrumental variable estimates for each exposure otherwise a random-effect model was used. All meta-analyses were performed in the R-based “meta” package (https://cran.r-project.org/web/packages/meta/index.html).

## Results

### Baseline characteristics of the study population

Of the 502,628 participants of the UK Biobank, a total of 288,572 were included in the final study sample for multivariable-adjusted survival analyses. Participants were excluded due to unavailable genetic data (N = 14,251), incomplete baseline data (N = 81,623), being related (N = 38,642), non-white British ancestry (N = 65,498) or unrealistic SD of autosomal probes (N = 9,440). 4,602 participants were excluded due to a history of stroke before study enrollment. The baseline characteristics of the participants divided over the quintiles of mtDNA abundance can be seen in **Table 1**. Participants in the lower lymphocyte mtDNA abundance quintiles had a higher mean age (57.5 versus 56.1 year), higher mean BMI (27.7 versus 27.0 kg/m^2^), were more frequently diagnosed with T2D (2.8% versus 2.0%) and were more frequently current smokers (11.3% versus 8.4%).

**Table 1.** Baseline characteristics of the study participants stratified by quintiles of mtDNA copy number.

|  | Q1 | Q2 | Q3 | Q4 | Q5 |
| --- | --- | --- | --- | --- | --- |
| N | 57 715 | 57 714 | 57 714 | 57 714 | 57 715 |
| Age (years) | 57.5 (8.0) | 57.1 (8.0) | 56.8 (8.0) | 56.5 (8.0) | 56.1 (8.0) |
| Sex (female %) | 52.0 | 53.2 | 54.0 | 54.3 | 55.7 |
| BMI (kg/m<sup>2</sup>) | 27.7 (5.0) | 27.5 (4.8) | 27.4 (4.7) | 27.2 (4.6) | 27.0 (4.5) |
| Diastolic blood pressure (mmHg) | 82.6 (10.2) | 82.4 (10.0) | 82.3 (10.0) | 82.1 (10.1) | 81.7 (10.1) |
| Systolic blood pressure (mmHg) | 139.5 (18.8) | 138.6 (18.7) | 138.2 (18.6) | 137.6 (18.3) | 136.7 (18.3) |
| Blood pressure-lowering medication % |  |  |  |  |  |
| Yes | 19.8 | 18.4 | 17.5 | 16.6 | 15.6 |
| No | 80.2 | 81.6 | 82.5 | 83.4 | 84.4 |
| Cholesterol (mmol/L) | 5.8 (1.2) | 5.7 (1.1) | 5.7 (1.1) | 5.7 (1.1) | 5.7 (1.1) |
| Cholesterol lowering medication % |  |  |  |  |  |
| Yes | 13.9 | 13.5 | 13.1 | 12.5 | 12.2 |
| No | 86.1 | 86.5 | 86.9 | 87.5 | 87.8 |
| Smoking % |  |  |  |  |  |
| Never | 53.3 | 54.1 | 55.0 | 55.6 | 56.5 |
| Past | 35.0 | 35.2 | 34.8 | 34.9 | 34.9 |
| Current | 11.3 | 10.4 | 9.9 | 9.4 | 8.4 |
| Type 2 Diabetes % |  |  |  |  |  |
| Yes | 2.8 | 2.5 | 2.3 | 2.2 | 2.0 |
| No | 97.2 | 97.5 | 97.7 | 97.8 | 98.0 |

Data are mean (SD) for continuous variables or percentages for dichotomous variables. Abbreviations: BMI, Body Mass Index.

### Multivariable-adjusted survival analyses lymphocyte mtDNA abundance and stroke

A total of 6,218 of the 288,572 participants (2.15%) had a stroke incidence, of which 3,994 (1.38%) were ischemic and 1,883 (0.65%) hemorrhagic during follow-up. The incidence of ischemic stroke was higher in the lower mtDNA abundance quintiles than in the higher quintiles in a step-wise manner (**Figure 1A**). Hemorrhagic stroke incidence was similar in all mtDNA abundance quintiles (**Figure 1B**).

**Figure 1.**
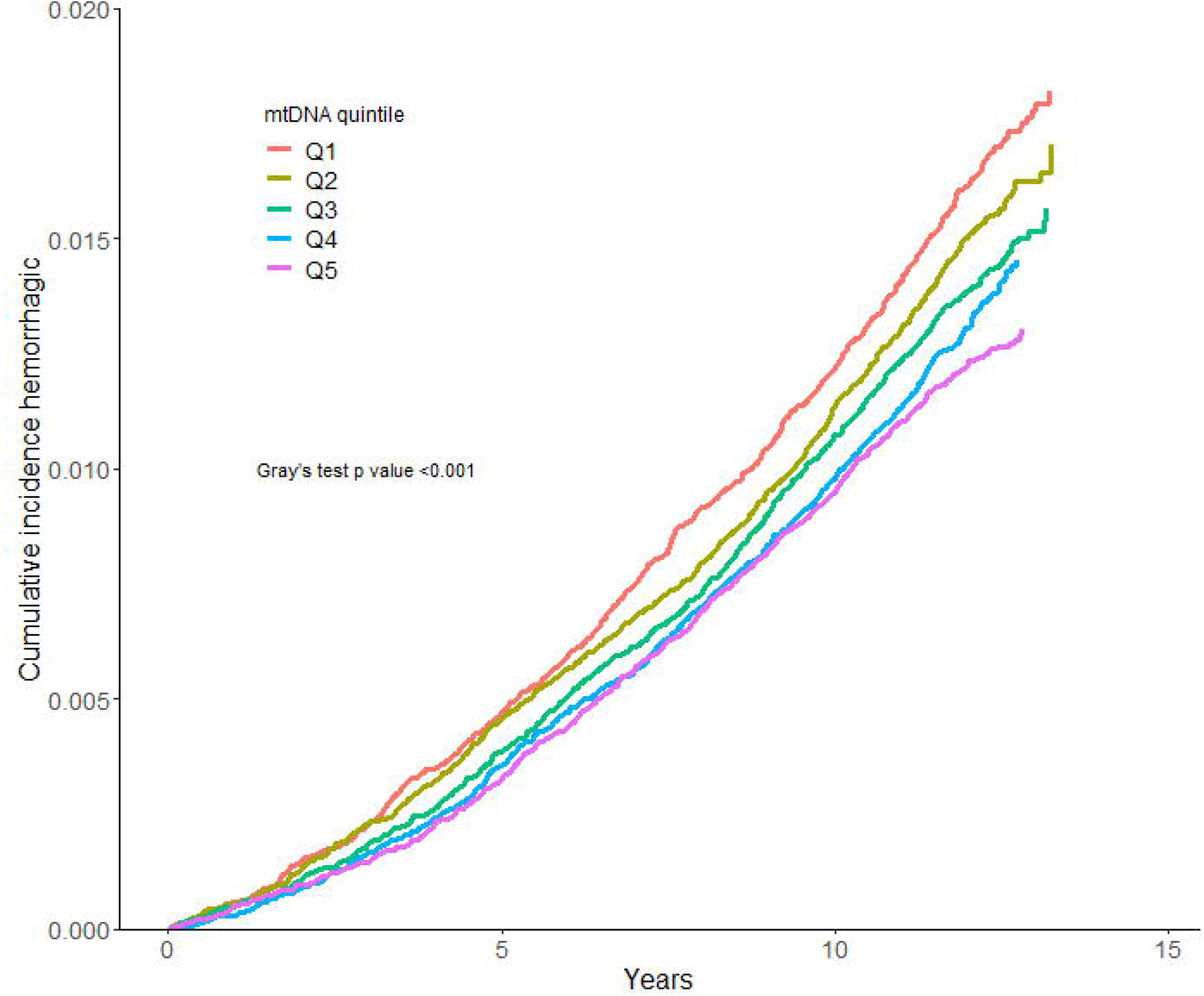
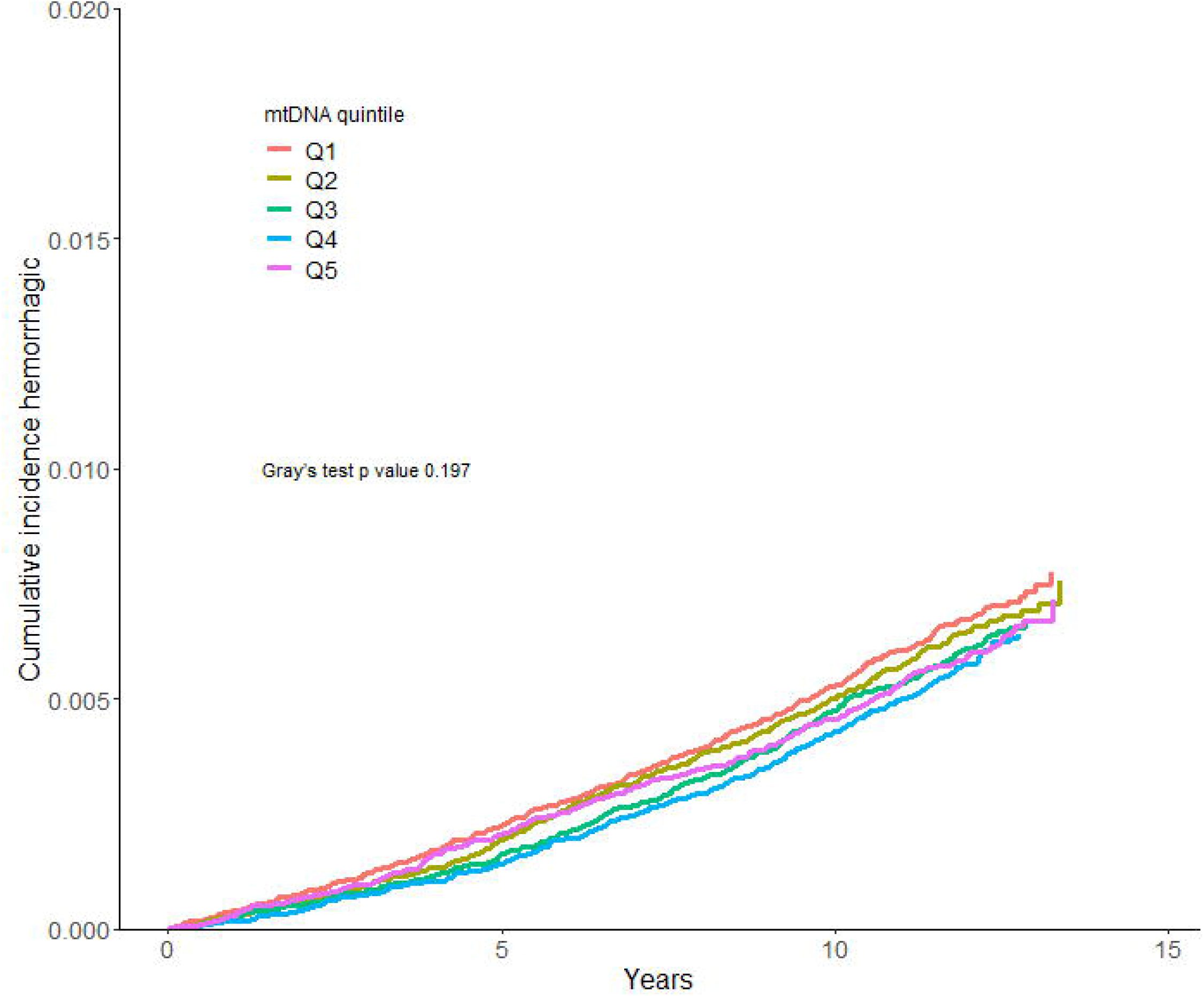
Cumulative incidence of ischemic (A) and hemorrhagic (B) stroke by quintiles of mtDNA copy number. We calculated the Cumulative incidence for ischemic and hemorrhagic stroke, accounting for death as a competing event. Differences in cumulative incidence between groups were assessed using Gray’s test.

**Table 2** shows the results for the association between mtDNA abundance and stroke incidence. After stratification, we found no evidence for effect modification by sex, so the data was treated as one sample and adjusted for sex. In model 1, mtDNA abundance was found to be associated with any stroke and ischemic stroke incidence, when comparing the first quintile with the highest 20% mtDNA abundance (ref) (any stroke: hazard ratio (HR), 1.10; 95% confidence interval (CI): 1.02 to 1.20; ischemic stroke: HR, 1.15; 95% CI: 1.04 to 1.27). No association was found between mtDNA abundance and incident hemorrhagic stroke (HR, 0.97; 95% CI: 0.84 to 1.13).

**Table 2.**
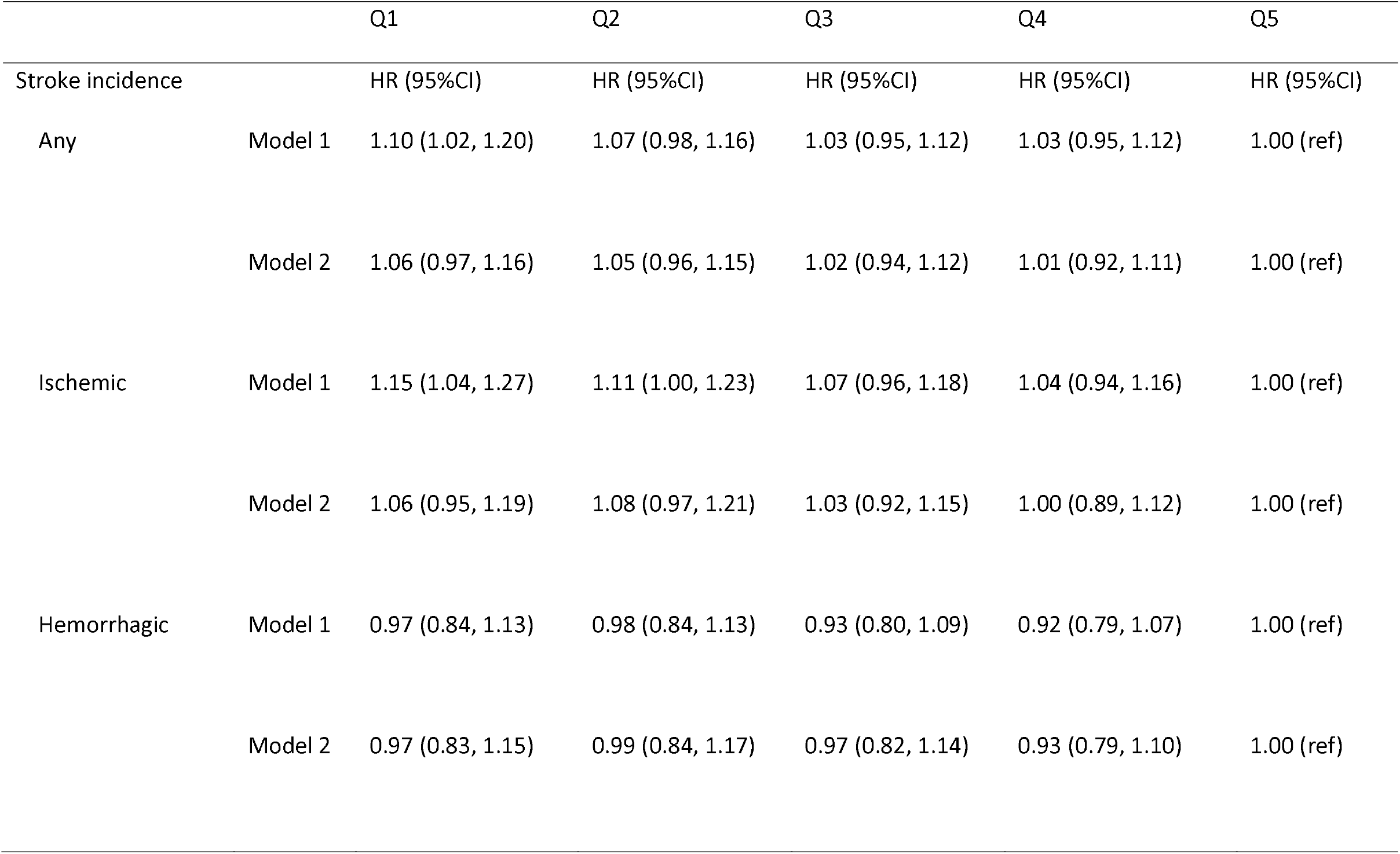
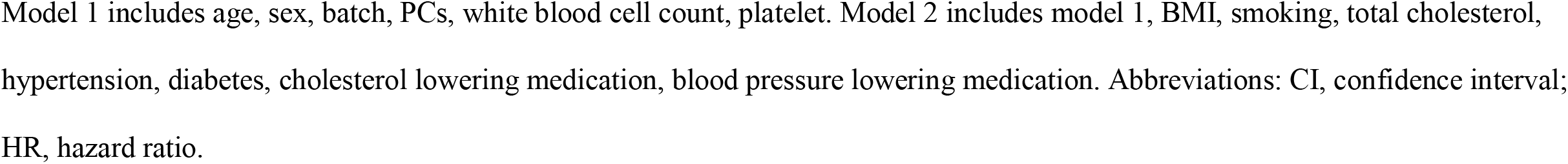
The multivariable-adjusted association between mtDNA copy numbers and incident stroke in European-ancestry participants from UK Biobank.

|  |  | Q1 | Q2 | Q3 | Q4 | Q5 |
| --- | --- | --- | --- | --- | --- | --- |
| Stroke incidence |  | HR (95%CI) | HR (95%CI) | HR (95%CI) | HR (95%CI) | HR (95%CI) |
| Any | Model 1 | 1.10 (1.02, 1.20) | 1.07 (0.98, 1.16) | 1.03 (0.95, 1.12) | 1.03 (0.95, 1.12) | 1.00 (ref) |
|  | Model 2 | 1.06 (0.97, 1.16) | 1.05 (0.96, 1.15) | 1.02 (0.94, 1.12) | 1.01 (0.92, 1.11) | 1.00 (ref) |
| Ischemic | Model 1 | 1.15 (1.04, 1.27) | 1.11 (1.00, 1.23) | 1.07 (0.96, 1.18) | 1.04 (0.94, 1.16) | 1.00 (ref) |
|  | Model 2 | 1.06 (0.95, 1.19) | 1.08 (0.97, 1.21) | 1.03 (0.92, 1.15) | 1.00 (0.89, 1.12) | 1.00 (ref) |
| Hemorrhagic | Model 1 | 0.97 (0.84, 1.13) | 0.98 (0.84, 1.13) | 0.93 (0.80, 1.09) | 0.92 (0.79, 1.07) | 1.00 (ref) |
|  | Model 2 | 0.97 (0.83, 1.15) | 0.99 (0.84, 1.17) | 0.97 (0.82, 1.14) | 0.93 (0.79, 1.10) | 1.00 (ref) |

Model 1 includes age, sex, batch, PCs, white blood cell count, platelet. Model 2 includes model 1, BMI, smoking, total cholesterol, hypertension, diabetes, cholesterol lowering medication, blood pressure lowering medication. Abbreviations: CI, confidence interval; HR, hazard ratio.

After correcting for BMI, smoking, alcohol consumption, total cholesterol, hypertension, diabetes, cholesterol lowering medication and blood pressure lowering medication, the associations found in model 1 attenuated (any stroke: HR, 1.06; 95% CI: 0.97 to 1.16; ischemic stroke: HR, 1.06; 95% CI: 0.95 to 1.19). In line with model 1, mtDNA abundance was not associated with hemorrhagic stroke when comparing groups after correction for other confounders (HR, 0.97; 95% CI: 0.83 to 1.17).

### Mendelian Randomization on mtDNA abundance and ischemic stroke

#### Main analyses

We did not observe any evidence for an association between genetically-influenced decrease in lymphocyte mtDNA abundance and ischemic stroke (**Figure 2**). The odds ratios (95% confidence intervals) per 1 SD less mtDNA-CN were 1.07 (95% CI: 0.95 to 1.20) in MEGASTROKE, 1.04 (95% CI: 0.79 to 1.37) in the UK Biobank, and 0.99 (95% CI: 0.82 to 1.20) in FinnGen. After meta-analysis, in a combined sample size of 1,098,740 (of which 73,296 cases), the pooled odds ratio was 1.04 (95%CI: 0.95 to 1.15) per 1-SD increase in genetically-influenced lymphocyte mtDNA abundance. Variance explained (R^2^) was 2.0% and calculated based on the derived summary statistics. MR-Egger, MR-PRESSO and weighted-median estimator regression were performed for each analysis separately and results were not materially different from the results observed with IVW, and hence no evidence for bias due to horizontal pleiotropy was identified.

**Figure 2.**
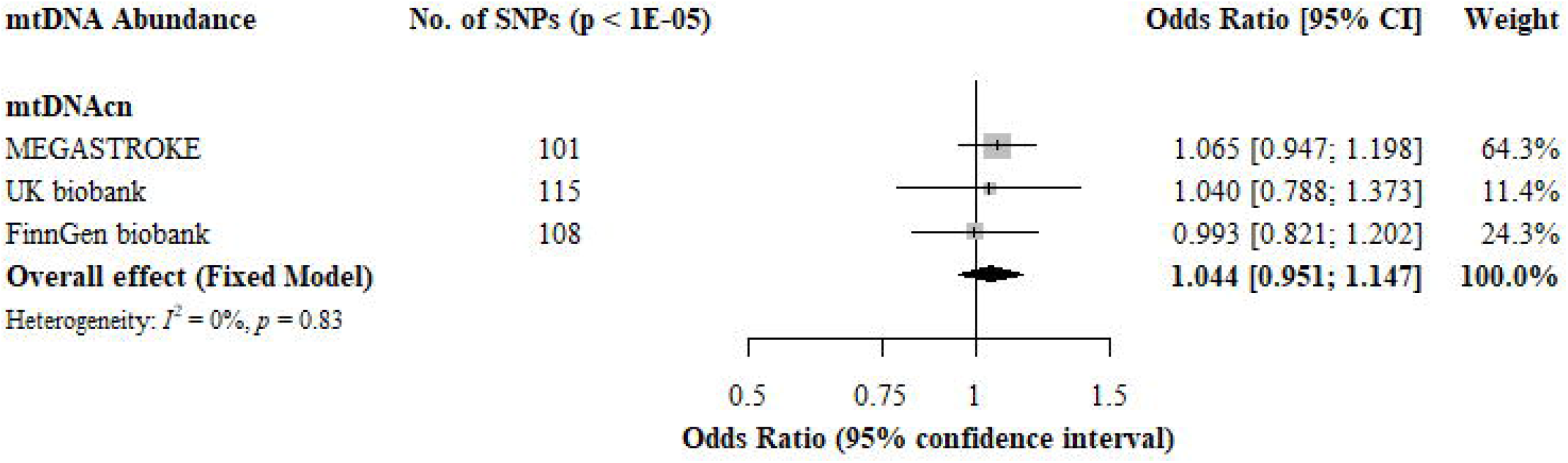
Causal association between mtDNA copy numbers and ischemic stroke occurrence. Estimated ORs represent the effect per SD decrease in mtDNA copy number on ischemic stroke. Results were opbtained using a Mendelian Randomization inverse-variance weighted method, analyzed per outcome database and combined over the three databases using fixed-effect meta-analyses.

#### Sensitivity analyses

A second MR analysis was performed using a different set of genetic variants for the exposure from *Hägg et al*.[10] Using these instruments, with an R^2^ of 1.3%, a 1-SD genetically-determined decrease in lymphocyte mtDNA abundance was not associated with a higher risk of ischemic stroke (OR: 1.05; 95% CI: 0.92 to 1.20), similar to the overall MR analysis (**Supplementary Figure 1**).

## Discussion

In our study, we investigated the association between lymphocyte mtDNA abundance as a proxy for mtDNA function,[8] and stroke incidence using both multivariable-adjusted survival analyses and MR approaches. In the UK Biobank cohort, consisting of 288,572 participants after exclusion, an initial association was found between mtDNA abundance and ischemic stroke. However, after correction for considered confounding factors the effect attenuated and was no longer significant. Thus, we did not find evidence of a relationship between mtDNA-CN and ischemic or hemorrhagic stroke occurrence risk using a prospective cohort study design. Consistent with the prospective analyses, the MR analyses, using a sample size of 73,296 cases and 1,025,444 controls, provided no evidence for an association between genetically-predicted mtDNA abundance and ischemic stroke. Collectively, these results indicate that there is no causal association between mtDNA abundance and stroke.

Several studies have shown an association between oxidative stress and secondary damage after stroke.[2, 3, 14] The biological explanation for this association was attributed to high concentrations of ROS leading to tissue destruction and cell death.[3] Our findings did not provide evidence for a relationship between oxidative stress and stroke occurrence, by analyzing lymphocyte mtDNA abundance as a marker of overall mitochondrial function.[8] We did find evidence of an association between mtDNA abundance and ischemic stroke after correction of only some technical variables. However, after correcting for considered confounders as BMI, diabetes, hypertension and total cholesterol, the association attenuated. This lead us to believe that the observed association between mtDNA abundance and ischemic stroke is confounded by these classic risk factors. In contrast to ischemic stroke we did not find an association between mtDNA abundance and hemorrhagic stroke in the univariate analyses. This difference might be explained because hemorrhagic stroke, in contrast to ischemic stroke, is also often caused by non-classic cardiovascular mechanisms such as vascular amyloid deposition in cerebral amyloid angiopathy.[25]

Earlier research has shown that obesity, diabetes and hypertension are associated with mitochondrial dysfunction.[26-30] Mitochondrial function has been associated with insulin sensitivity, explaining the link with type 2 diabetes.[31] However, the authors state that it remains a topic of discussion whether mitochondrial dysfunction is a cause or consequence of diabetes pathology. When looking specifically at the cardiovascular system, it has been reported that ROS is increased in the diabetic heart.[32] The relationship between cholesterol and mitochondrial dysfunction is far less well understood. The few studies that have been done on this topic focus mainly on the metabolic syndrome, which is characterized by a combination of increased cardiometabolic risk factors. These studies indicate that the relationship between mitochondria and the pathogenesis of the metabolic syndrome is yet to be completely unraveled. (reviewed in [33]) In combination with our findings, these studies might implicate a relationship between mitochondrial dysfunction and ischemic stroke only when confounded by these primary risk factors.

Our data on mtDNA abundance was obtained from lymphocytes. Although some of the lymphocytes may be directly involved in the pathology of stroke, additional cell types such as endothelial and smooth muscle cells, that we did not query for mitochondrial abundance, are clearly more directly involved. This could potentially explain our null findings. Studies on the difference in mitochondrial function within an individual between cell groups are, to our knowledge, non-existent. The evidence thus far indicates that mitochondrial dysfunction is systemic as, for example, the consequences of reduced mitochondrial function -reduced insulin sensitivity-can also be found in the brain.[31]

A key strength of this study is the statistical power of the analyses of the association between stroke and mitochondrial abundance (288,572 participants for the multivariable survival analysis and 1,098,740 for the MR, respectively). In addition, the use of multivariable survival analysis and MR, with comparable results all point towards the same conclusion. We were able to adjust for some of the primary risk factors on stroke, making it probable that any effect found could be attributed to our exposure. In line, finding no effect after correction indicates that lymphocyte mtDNA abundance has no association with stroke in our study population.

Some limitations are to be considered. First, our study population consists of predominantly Caucasian participants, limiting the generalizability of the results to other ancestry groups. Second, no information on the underlying causes of stroke was available in the UK biobank. Stroke is a heterogenous disease and we cannot exclude that the role of mitochondrial dysfunction and stroke occurrence differs for different stroke subtypes. Last, despite a large sample size in the multivariable adjusted analysis, stroke and especially hemorrhagic stroke incidences were relatively few. However, as an association was found before correction, we think our analyses had enough power to detect a difference between groups. In addition, we used one of the larger data sets available. It is therefore most likely that the association between mtDNA-CN and ischemic stroke is confounded by classic (cardiovascular) risk factors.

In conclusion, despite a large sample size our study did not find evidence for a causal association between mtDNA abundance and ischemic or hemorrhagic stroke. It is therefore likely that mtDNA abundance is a marker of ischemic, not hemorrhagic, stroke risk but not a direct causal player.

## Data Availability

All data produced in the present study are available upon reasonable request to the authors

## Abbreviations

GWAS: Genome Wide Association Study
HR: Hazard Ratio
MR: Mendelian Randomization
mtDNA-CN: mitochondrial DNA Copy Numbers
OR: Odds Ratio
ROS: Reactive Oxygen Species
SNP: Single-Nucleotide Polymorphisms

## Acknowledgements

The authors are grateful to the UK Biobank for allowing us the use of their data. The analyses done in UK Biobank were done under project number 56340. Furthermore, the authors acknowledge the participants and investigators of the MEGASTROKE consortium and the FinnGen Biobank who contributed to the summary statistics data which are made available for further studies.

LGM, JL, RN and DvH designed research; LGM and JL conducted research; LGM and JL performed statistical analysis; LGM, JL, RN, and MJHW wrote paper; LGM had primary responsibility for final content. DvH, KWvD, SH, FG and MJHW contributed to the data interpretation and commented on initial versions of the manuscript; All authors read and approved the final manuscript.

