## Supplementary figures and images for "The association between lymphocyte mitochondrial DNA abundance and Stroke: a combination of multivariable-adjusted survival and mendelian randomization analyses"

### Supplemental Figure 1

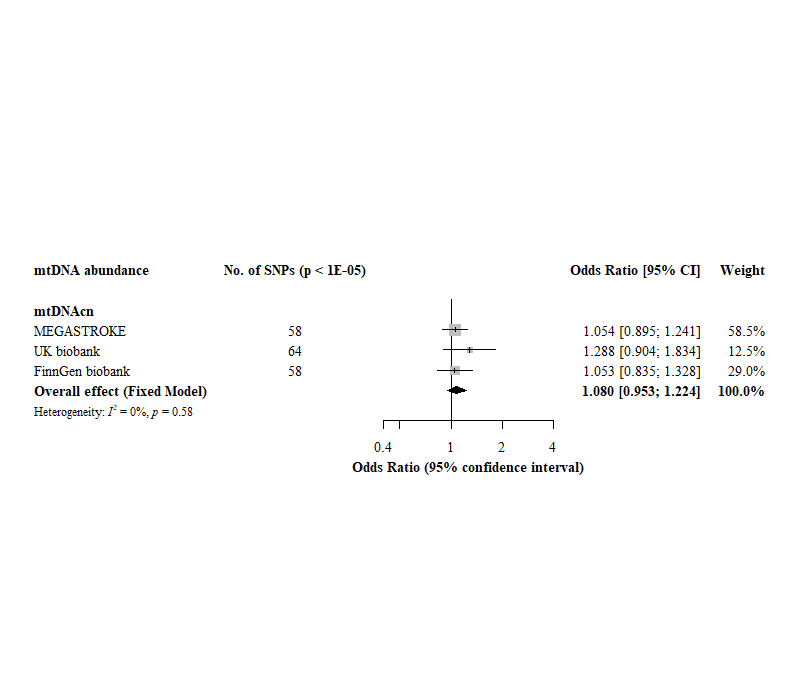
